# Use of Artificial Intelligence for Acquisition of Limited Echocardiograms: A Randomized Controlled Trial for Educational Outcomes

**DOI:** 10.1101/2023.04.12.23288497

**Authors:** Evan Baum, Megha D. Tandel, Casey Ren, Yingjie Weng, Matthew Pascucci, John Kugler, Kathryn Cardoza, Andre Kumar

**Affiliations:** Department of Medicine, Stanford University School of Medicine, Stanford, CA; Quantitative Sciences Unit, Department of Medicine, Stanford University School of Medicine, Stanford, CA; Department of Medicine, Cedars Sinai, Los Angeles, CA

**Keywords:** bedside ultrasound, point-of-care ultrasound, competency, technique, patient care, medical education, echocardiogram

## Abstract

**Background:** Point-of-care ultrasound (POCUS) machines may utilize artificial intelligence (AI) to enhance image interpretation and acquisition. This study investigates whether AI-enabled devices improve competency among POCUS novices.

**Methods:** We conducted a randomized controlled trial at a single academic institution from 2021-2022. Internal medicine trainees (N=43) with limited POCUS experience were randomized to receive a POCUS device with (Echonous, N=22) or without (Butterfly, N=21) AI-functionality for two weeks while on an inpatient rotation. The AI-device provided automatic labeling of cardiac structures, guidance for optimal probe placement to acquire cardiac views, and ejection fraction estimations. Participants were allowed to use the devices at their discretion for patient-related care.

The primary outcome was the time to acquire an apical 4-chamber (A4C) image. Secondary outcomes included A4C image quality using the modified Rapid Assessment for Competency in Echocardiography (RACE) scale, correct identification of pathology, and participant attitudes. Measurements were performed at the time of randomization and at two-week follow-up. All scanning assessments were performed on the same standardized patient.

**Results:** Both AI and non-AI groups had similar scan times and image quality scores at baseline. At follow-up, the AI group had faster scan times (72 seconds [IQR 38-85] vs. 85 seconds [IQR 54-166]; p=0.01), higher image quality scores (4.5 [IQR 2-5.5] vs. 2 [IQR 1-3]; p<0.01) and correctly identified reduced systolic function more often (85% vs 50%; p=0.02) compared to the non-AI group. Trust in the AI features did not differ between the groups pre- or post-intervention. The AI group did not report increased confidence in their abilities to obtain or interpret cardiac images.

**Conclusions:** POCUS devices with AI features may improve image acquisition and interpretation by novices. Future studies are needed to determine the extent that AI impacts POCUS learning.

## Introduction

Point-of-care ultrasonography (POCUS) describes the use of ultrasound by clinicians at the bedside to provide real-time diagnoses and assist with procedural interventions.^1^ It has been shown to improve diagnostic accuracy, reduce procedural complications, reduce direct costs for healthcare organizations, and improve patient satisfaction by encouraging the clinician to be present at the bedside.^1–5^

In response to the growing evidence supporting the use of POCUS for patient care, medical schools, residency programs, and professional societies have developed training programs for its safe and effective usage.^5–7^ In addition, several organizations have established guidelines in an effort to standardize POCUS training.^3,8^ Barriers to implementing more universal training measures include time constraints within training programs, a paucity of faculty credentialed for supervision, limiting funding, the need for established quality assurance protocols, and a lack of standardized assessments.^1,6,7,9^ Due to these barriers, novice users continue to use POCUS with minimal training and oversight,^5,10^ thus underscoring the need for alternative training methods with this ever-growing technology.

POCUS device manufacturers have begun to employ artificial intelligence (AI) to aid in image acquisition and interpretation, which may help address the current training barriers faced by clinicians.^11^ This technology could become complementary to traditional teaching methods by offering augmented guidance of probe placement to improve a user’s manual dexterity and labeled images to aid in the rapid identification of pathology.^12^ Previous investigations have shown that AI-assisted ultrasounds can aid novices in obtaining high quality cardiac images, accurately assessing ventricular function, and identifying non-trivial pericardial effusions.^13,14^ One prospective study among emergency medicine trainees demonstrated that AI-augmented POCUS may increase the efficiency and accuracy of diagnosing pneumonia in pediatric populations.^15^ There are currently no randomized studies comparing POCUS image acquisition and interpretation with AI vs. non-AI equipped devices among inexperienced users.

This randomized, controlled trial aims to address several ongoing gaps in knowledge related to POCUS learning. First, we hypothesize that POCUS novices randomized to AI-enabled devices that aid in image acquisition and interpretation will be more efficient and proficient at these tasks than those without AI-enabled devices. Secondly, we hypothesize that those who used the AI-devices will feel more confident in their abilities due to the additional assistance this technology may provide during scanning.

## Methods

### Study Participants & Setting

We conducted a randomized controlled trial at a single academic institution from 6/2021-1/2022. All eligible residents were recruited via email explaining the general nature of the study. Our inclusion criteria included internal medicine residents rotating on the general inpatient wards service. We excluded residents who had taken an ultrasound elective offered by our residency program. At the time of the study, there was no formal POCUS credentialing pathway within the residency program. The Stanford University Institutional Review Board approved this investigation.

### Study Design

Recruited residents (N=43) were randomized 1:1 to receive a POCUS device with AI-functionality (Echonous, N=22) or without (Butterfly, N=21) for two weeks (Figure 1). Outcomes were assessed at baseline and at two-week follow-up (see Outcomes below). Participants were allowed to use the devices at their discretion for patient-related care or self-directed learning. For privacy reasons, any saved patient images were not reviewed by the researchers and feedback was not provided on participants’ scans. At the time of randomization, all participants received written and verbal instructions on their device’s functionality and access to online learning modules regarding POCUS acquisition and interpretation (Appendix).

**Figure 1.**
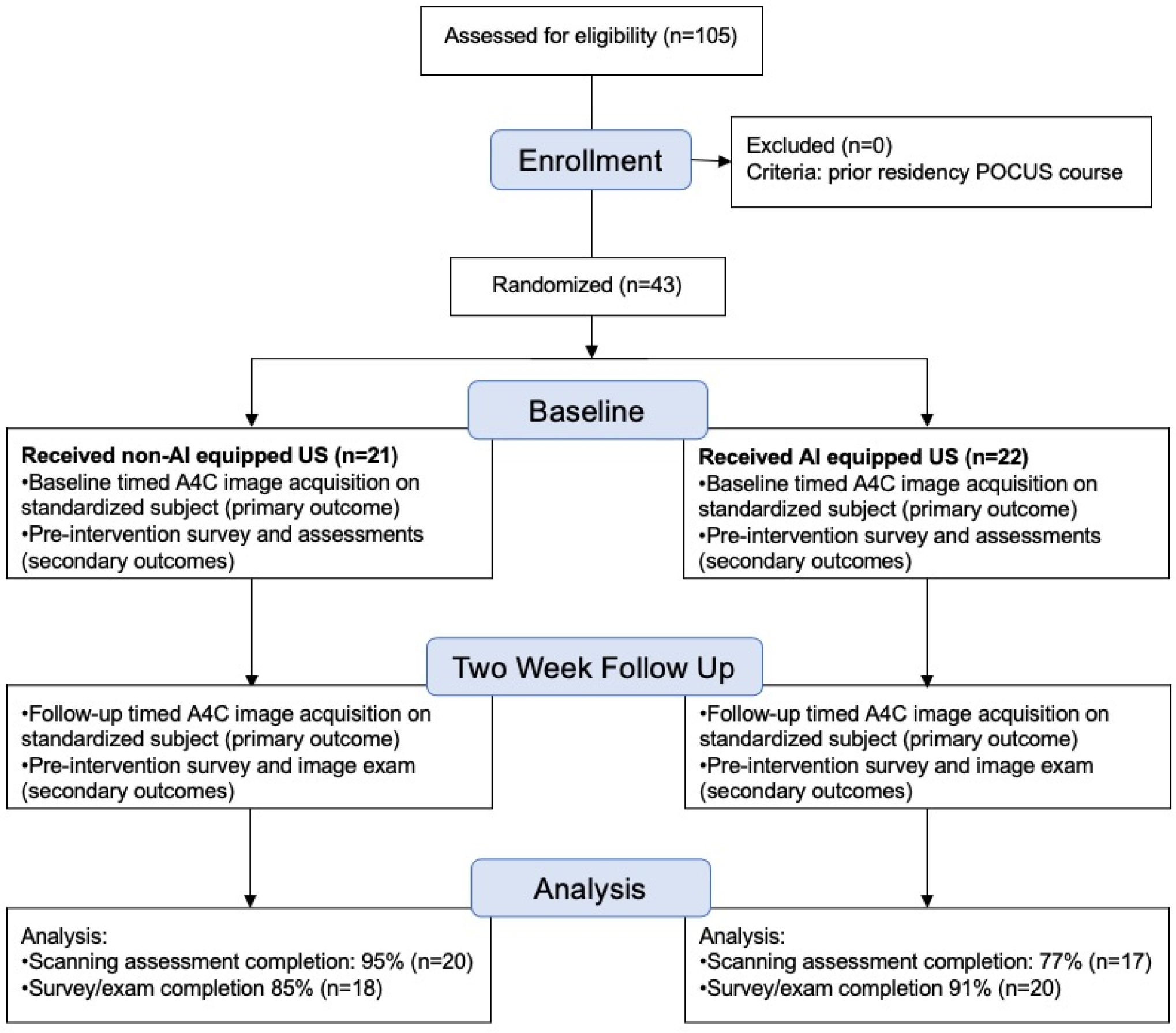
Overview of Study. A4C, apical-4-chamber view.

Participants were instructed to use an electronic log each time they used their devices for any type of scan to track how frequently they were being used.

### Devices

Two handheld POCUS devices were utilized in this study: Echonous^™^ (AI) and Butterfly^™^ (non-AI). These handheld devices were chosen for this study due to budgetary constraints and the unavailability of other machines. The AI device provided automatic labeling of anatomic structures, real-time guidance for optimal probe placement to acquire an apical-4-chamber view (A4C), and automatic left ventricular systolic function estimation using the apical windows (Appendix). The non-AI device did not provide these features (Appendix).

### Outcomes

Our primary outcome was the time to acquire an A4C image. Secondary outcomes included the quality of captured A4C images, correct interpretation of pathological images, correct identification of anatomic structures, trainee confidence in POCUS, and their trust in the AI system.

### Assessments/Surveys

Measurements were performed at the time of randomization (baseline) and at two-week follow-up. All of the scanning assessments were performed on the same standardized patient with the same probe (Butterfly), regardless of study arm. A study author was present for the scanning assessments to provide instruction and to set up the device, but they did not directly observe or comment on the images being acquired. For the primary outcome of scanning time, participants were instructed to notify the proctor when they had acquired an optimal A4C image for evaluation (which was saved for analysis). Participants were timed from the moment the probe touched the patient’s torso until proctor notification. For the secondary outcome of scan quality, we utilized the modified Rapid Assessment of Competency in Echocardiography (RACE) scale, which has excellent interrater reliability (α = 0.87) and has been previously validated as an assessment tool for image acquisition and quality with POCUS.^16,17,16,18^ Two reviewers (AK and JK), who were blinded to the study arms, independently reviewed the baseline and follow-up assessment scans to assign RACE scores. The average scores between the two reviewers were used to create composite RACE scores for analyses.

Assessments of anatomic identification and pathological image interpretation were performed utilizing a HIPPA-compliant online survey platform (Qualtrics, Provo, UT) that was sent pre- and post-intervention. This assessment has been previously described by our study team and consists of short video clips with multiple choice answers.^5^ Surveys were administered alongside these assessments to the participants. These surveys assessed trainee attitudes toward POCUS, trust in the AI system, and their own confidence in acquiring images. Attitudes were measured using 5-point Likert scales. These surveys were based on previously described assessments.^5,10^

### Statistical Analysis

Baseline performance and attitudes were compared between participants in the AI group and those in the non-AI group.

Distributions of the scanning time were visualized in a Box/violin plot. Median and interquartile range (IQR) were reported and compared by the two randomized groups. We further performed the ANOVA (or Kruskal-Wallis) tests to compare the distributions by the two randomized groups at two-week follow-up visits. Similar analysis was performed for all secondary outcomes. To explore the potential changes of the outcomes over time, Wilcoxon signed rank test was applied to compare the differences of secondary outcomes from baseline to 2-week follow-up, by the two randomized groups, respectively. Median and IQR were reported for baseline and 2-week follow-up. Chi-square tests were performed to compare participants’ confidence levels.

All statistical tests were conducted using SAS 9.4 (Cary, NC), and a p-value <0.05 was considered statistically significant.

## Results

### Baseline Characteristics

There were a total of N=105 residents eligible for participation, of which N=43 responded to the email invitation to participate. No residents were excluded. Among the N=43 residents, N=22 were randomized to the AI device, while N=21 were randomized to the non-AI device.

Completion rates at follow-up for the scanning assessments were 77% (N=17) for the AI group and 95% (N=20) for the non-AI group (Table 1). Survey and image quiz completion rates at follow-up were 91% (N=20) for the AI group and 85% (N=18) for the non-AI group. Participant demographics and their prior POCUS experience are shown in Table 1.

**Table 1.** Participant Demographics and Completion Rates. AI, artificial intelligence; PGY, post-graduate year.

|  | Baseline AI | Baseline Non-AI | Post-Intervention AI | Post-Intervention Non-AI |
| --- | --- | --- | --- | --- |
| Completed Scanning Assessments (%) | 22 (100%) | 21 (100%) | 17 (77%) | 20 (95%) |
| Completed Surveys & Quiz (%) | 22 (100%) | 21 (100%) | 20 (91%) | 18 (85%) |
| PGY-1 (%) | 6 (27%) | 6 (29%) | 6 (30%) | 6 (33%) |
| PGY-2 (%) | 11 (50%) | 12 (57%) | 11 (55%) | 10 (56%) |
| PGY-3 (%) | 5 (23%) | 3 (14%) | 3 (15%) | 2 (11%) |
| Females (%) | 10 (45%) | 8 (38%) | 9 (45%) | 6 (33%) |
| Exposure to Prior POCUS Courses (%) | 3 (14%) | 3 (14%) | 2 (10%) | 3 (17%) |

### Primary Outcome: Time to Scan

For our primary outcome of time to acquire an A4C image at follow-up, the scanning times were significantly faster in the AI (median 57s [IQR: 32-75]) vs. non-AI groups (median 85.0s [IQR: 50-172]; p=0.01; Table 2). Both groups had similar median scanning times at baseline (AI 146s [IQR: 98-220] and non-AI 119s [IQR: 64-175]; p=0.21). On sub-analysis, the AI group significantly improved in their median scanning times pre-vs. post-intervention (p<0.01), while the non-AI group did not (p=0.26).

**Table 2.**
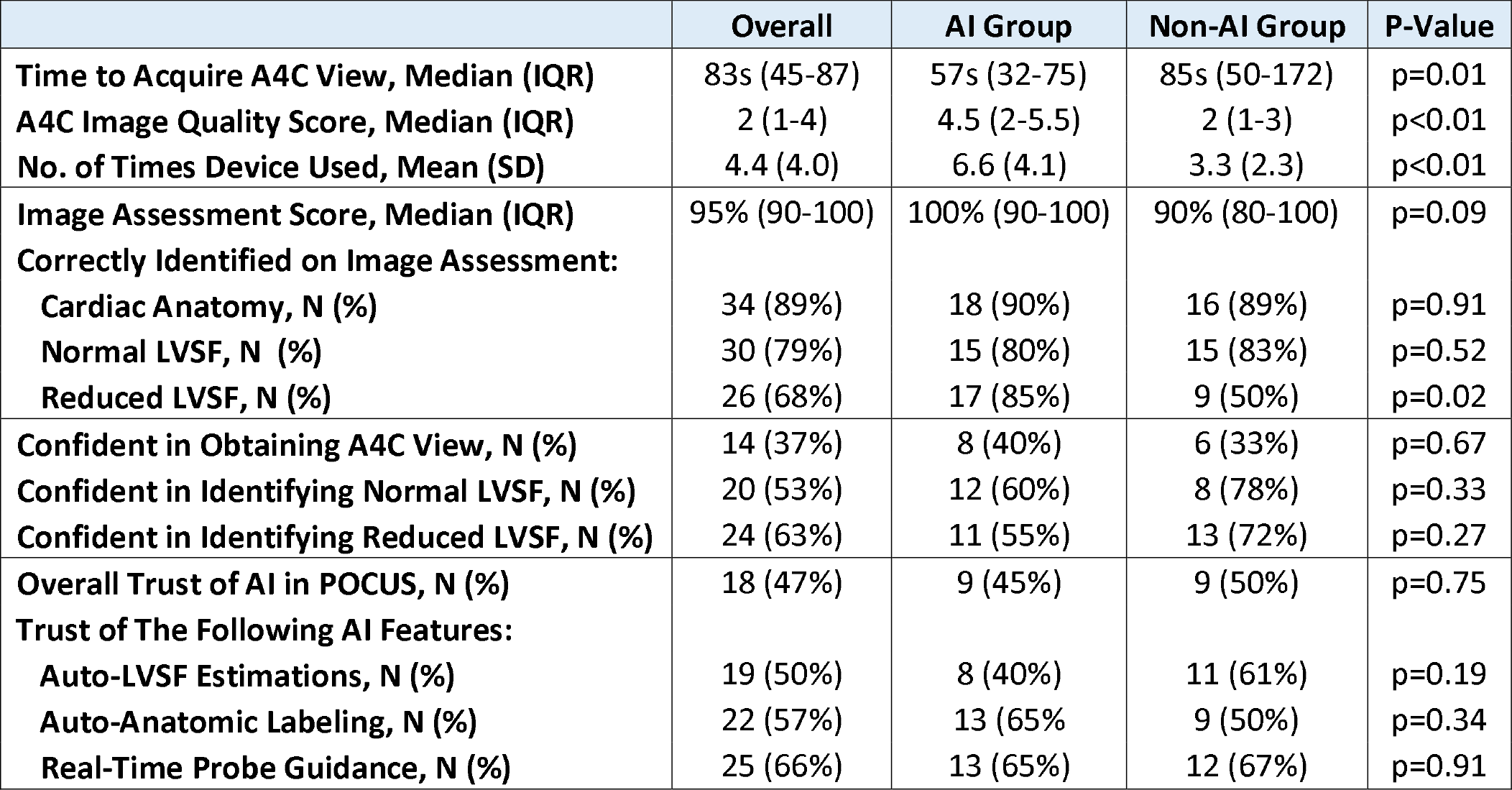
Study Outcomes at Two-Week Follow-Up. A4C, Apical 4-Chamber View; IQR, Interquartile Range; LVSF, Left-Ventricular Systolic Function.

|  | Overall | AI Group | Non-AI Group | P-Value |
| --- | --- | --- | --- | --- |
| Time to Acquire A4C View, Median (IQR) | 83s (45-87) | 57s (32-75) | 85s (50-172) | p=0.01 |
| A4C Image Quality Score, Median (IQR) | 2 (1-4) | 4.5 (2-5.5) | 2 (1-3) | p<0.01 |
| No. of Times Device Used, Mean (SD) | 4.4 (4.0) | 6.6 (4.1) | 3.3 (2.3) | p<0.01 |
| Image Assessment Score, Median (IQR) | 95% (90-100) | 100% (90-100) | 90% (80-100) | p=0.09 |
| Correctly Identified on Image Assessment: |  |  |  |  |
| Cardiac Anatomy, N (%) | 34 (89%) | 18 (90%) | 16 (89%) | p=0.91 |
| Normal LVSF, N (%) | 30 (79%) | 15 (80%) | 15 (83%) | p=0.52 |
| Reduced LVSF, N (%) | 26 (68%) | 17 (85%) | 9 (50%) | p=0.02 |
| Confident in Obtaining A4C View, N (%) | 14 (37%) | 8 (40%) | 6 (33%) | p=0.67 |
| Confident in Identifying Normal LVSF, N (%) | 20 (53%) | 12 (60%) | 8 (78%) | p=0.33 |
| Confident in Identifying Reduced LVSF, N (%) | 24 (63%) | 11 (55%) | 13 (72%) | p=0.27 |
| Overall Trust of AI in POCUS, N (%) | 18 (47%) | 9 (45%) | 9 (50%) | p=0.75 |
| Trust of The Following AI Features: |  |  |  |  |
| Auto-LVSF Estimations, N (%) | 19 (50%) | 8 (40%) | 11 (61%) | p=0.19 |
| Auto-Anatomic Labeling, N (%) | 22 (57%) | 13 (65%) | 9 (50%) | p=0.34 |
| Real-Time Probe Guidance, N (%) | 25 (66%) | 13 (65%) | 12 (67%) | p=0.91 |

### Secondary Outcomes

#### A. Image Quality

For the secondary outcome of A4C image quality at follow-up, the median RACE scores were significantly higher in the AI (4.5 points [IQR 2-5.5]) vs. the non-AI group (2 points [IQR 1-3]; p<0.01; Table 2). Both groups had statistically similar median RACE scores at baseline (AI: 3 points [IQR 2-4]; non-AI: 2 points [IQR: 1-3]; p=0.08).

#### B. Identification of Pathology and Anatomy

Overall, the AI and non-AI groups performed similarly on the follow-up image assessment for left ventricular systolic function and anatomic identification in the A4C view (AI median score: 100% [IQR: 90-100%] vs. non-AI median score: 90% [IQR: 80-100%]; p=0.09; Table 2). Notably, a greater proportion of the AI group correctly identified reduced left ventricular systolic function in the A4C view compared to the non-AI group on the two-week follow-up assessment (85% vs. 50%; p=0.02).

#### C. Device Usage

Participants in both arms were tracked on the frequency they used the devices using an electronic log (see Methods). Participants randomized to the AI device reported using the devices nearly twice as frequently as those randomized to a non-AI device (mean 6.6 times [SD 4.1] vs. 3.3 times [SD 2.3] ; p<0.01; Table 2).

#### D. Survey Results

The AI and non-AI participants reported similar confidence levels being able to obtain A4C images at two-week follow-up (Table 2), despite the AI group having significantly faster scan times and higher image quality scores. Similarly, both groups reported similar confidence in identifying normal and reduced LV systolic function (Table 2). When the pre-vs. post-intervention surveys for the AI group were compared, they did not report an increase in trust for the AI features (anatomic labeling, calculations of ejection fraction, and real-time guidance for optimal probe placement; Appendix). Both groups reported low to moderate trust in the AI system on the post-intervention surveys, including for features directly exhibited by the AI device (Table 2).

## Discussion

As POCUS usage continues to expand,^7,19^ AI-enabled devices represent a possible means to enhance competency among novice users and aid in interpretation.^13,14,20^ In this randomized, controlled trial, we observed that internal medicine residents randomized to carry an AI-POCUS device for two weeks without feedback were able to obtain A4C views more quickly, had higher A4C image quality scores, and were more likely to identify reduced systolic function compared to residents who carried non-AI devices. Interestingly, the general trust in the AI system remained low to moderate in both groups, and the AI group did not report higher confidence in their skills despite outperforming the non-AI group. To our knowledge, this is the first randomized study that demonstrates that AI can improve scanning efficiency, acquired image quality, and pathological image interpretation.

The use of AI-enabled technologies to enhance proficiency in performance is gaining attention as an alternative to traditional teaching methods.^21–23^ Previous non-randomized studies have shown that AI-assisted ultrasounds can aid in the acquisition of cardiac images and interpretation of reduced systolic function.^13,14^ While our results support the hypothesis that AI-enabled devices can improve POCUS learning among novices, it is important to note the AI group in this study performed more scans overall. It could be argued this alone led to more deliberate practice and improved competency in the AI group. In support of this, previous investigations have shown that trainees can become proficient in acquiring cardiac and abdominal POCUS images in as few as 20-30 examinations.^16,18,24,25^ However, it is important to consider whether such trainees would be considered “competent” with POCUS.^18^ Some authors have argued that the mastery of skills requiring manual dexterity may take substantially longer and require years of deliberate practice.^26,27^ Furthermore, others have demonstrated that the degree of improvement for cardiac images is nominal between 0-10 scans (our groups had a mean difference of roughly three scans).^16,17,16,18^

In this study, we found the participants had low to moderate levels of trust in the AI system, and the AI-group did not report increased trust in the system at follow-up. This lack of trust despite improved performance with AI is well described outside of POCUS,^22,28,29^ which underscores the need for effective curricula on how to optimally integrate AI with clinical care.^30^ Moreover, the AI group did not report higher confidence levels in their own skills despite significantly higher levels of performance. This finding is consistent with previous investigations that have demonstrated a novice’s actual ability to acquire ultrasound images does not correlate with their expressed confidence.^10^ Our results reinforce a concern that novices may have difficulty assessing their own competency with POCUS, which underscores the need for stringent training requirements and oversight as the technology and its usage expands.^18,31,32^

There are several limitations of this study. It was conducted at a single academic site, which limits its generalizability. Due to the study design, participants were not blinded to the study outcomes or their study arm. Moreover, the relatively short-term follow up rate limits any conclusions regarding skill retention or overall improvement in competency. No formal feedback was provided to either study arm on image quality or acquisition techniques, even though this is thought to be an effective means of teaching POCUS.^6^ The scanning assessments were performed on a standardized patient with adequate cardiac windows and the image assessments were performed using idealized POCUS images. Therefore, these findings may not reflect real world practice wherein clinicians obtain images and evaluate them at bedside, often in patients with difficult anatomy. Nevertheless, these results represent an intriguing implementation of AI-enabled POCUS, with future studies being warranted to investigate its applications in medical training and cardiac image acquisition.

In conclusion, POCUS novices randomized to carry AI-enabled devices for two weeks for patient care were able to obtain cardiac images more quickly, had higher image quality scores, and more accurately identified reduced systolic function at two week-follow-up. However, they continued to have low to moderate trust in the device’s AI features despite superior performance. Future studies should focus on how AI impacts long-term POCUS learning and the retention of skills.

## Data Availability

ll data produced in the present study are available upon reasonable request to the authors

## Abbreviations

A4C: (Apical-4-chamber)
AI: (artificial intelligence)
POCUS: (point-of-care ultrasound)
RACE: (rapid assessment for competency in echocardiography)

## Acknowledgements

None

