## Appendix Material for "Use of Artificial Intelligence for Acquisition of Limited Echocardiograms: A Randomized Controlled Trial for Educational Outcomes"

**Supplementary Material**

**Appendix A:** Link to online learning modules used for this study: <https://stanforduniversity.qualtrics.com/jfe/form/SV_8IKCWPdttGhM8Yd>

**Appendix B:** Link to survey and image examinations used for this study:

<https://stanforduniversity.qualtrics.com/jfe/form/SV_410pQUZfXOH2Nka>

**Appendix C:**

Written Materials Given for AI Device Functionality: <https://cdn-echonous.b-cdn.net/wp-content/uploads/2023/02/P005794-023_-Rev-A_-Kosmos_User-Guide_-v7.2_clean-1.pdf>

Written Materials Given for Non-AI Device Functionality: https://manual.butterflynetwork.com/butterfly-iq-user-manual_rev-bc-en.pdf

**Appendix D:** Copy of the Modified RACE (Rapid Assessment of Competency in Echocardiography) rubric

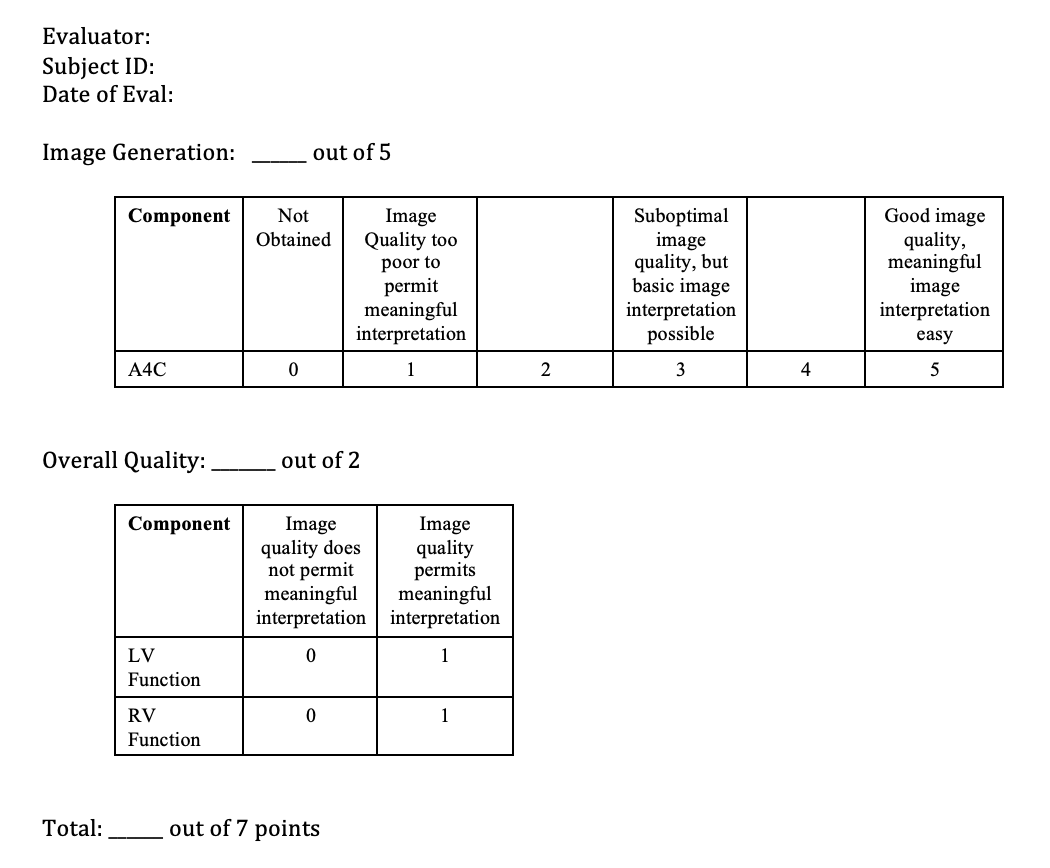

Appendix E. Comparison of Pre- vs. Post-Intervention Attitudes Among the AI-Participants. Analysis was performed using Chi-Square analysis. AI, artificial intelligence; LVSF, left ventricular systolic function.

|  | **AI Pre-Intervention (N=22)** | **AI Post-Intervention (N=20)** | **p-value** |
| --- | --- | --- | --- |
| **Overall Trust of AI in POCUS, N (%)** | 6 (27%) | 9 (45%) | 0.21 |
| **Auto-LVSF Estimations, N (%)** | 7 (32%) | 8 (40%) | 0.58 |
| **Auto-Anatomic Labeling, N (%)** | 9 (41%) | 13 (65%) | 0.12 |
| **Real-Time Probe Guidance, N (%)** | 12 (54%) | 13 (65%) | 0.48 |
